# Efficacy of recombinant human epidermal growth factor (rhEGF) incorporated in an absorbable collagen membrane for the management of gingival recession defects

**DOI:** 10.1101/2023.01.25.23284988

**Authors:** Rampalli Viswa Chandra, Kotha Sushmitha Bindu, Aileni Amarendhar Reddy

**Affiliations:** Department of Periodontics, SVS Institute of Dental Sciences, Yedira Post, Mahabubnagar, Telangana, India-509002

**Keywords:** Epidermal growth factor, Growth factor, Gingival recession

## Abstract

**Aim & Objectives:** The aim of the present study is to evaluate the regeneration efficacy of rhEGF impregnated in collagen membrane for the management of Millers class I & class II gingival recession defects.

**Patients and methods:** 18 patients with 30 Millers class I & class II gingival recession defects were treated with one of the following interventions and randomly allocated into each of the following experimental groups; Test group: rhEGF impregnated in collagen membrane, Control group: plain collagen membrane. Clinical measurements at baseline, 3 months and 6 months included decreased probing depth, recession depth and increase in width of keratinized gingiva.

**Results:** There was an improvement in tissue biotype in test group and statistically significant increase in KGW from baseline to 3 months which remained constant from 3 months to 6 months(p≤0.001) in both the groups. Similarly, RD shows constant increase from baseline to 6 months in test group whereas there is reduction in control group (p≤0.003). There was significant difference in clinical parameters in both test and control groups. All the patients had an uneventful healing phase.

**Conclusion:** The beneficial effects of rhEGF resulted in healthy wound healing process with less scarring offers more potential properties showed promising results over collagen membrane. Further larger samples are required to confirm the efficacy of rhEGF in root coverage for soft tissue regeneration.

## INTRODUCTION

Apical migration of the marginal gingiva in relation to the cemento-enamel junction, gingival recession exposes the tooth’s root surface.^[1]^ As root exposure is frequently linked to dentinal hypersensitivity, esthetic concerns and root cavities, increased incidence of recession has become a key concern for clinicians. A number of predisposing variables, including the eruption pathway, malposition, muscle attachment, frenal pull, alveolar bone dehiscence etc., are linked to marginal gingival recession.^[2]^ In addition to these risk factors, the thin gingival biotype—where there is a delicate, thin marginal tissue covering the avascular root surface—is the most prevalent and significant risk factor.^[3]^ Chronic mechanical or frictional damage, biofilm buildup, a small band of keratinized mucosa or both are usually linked to single or multiple recessions.

A number of surgical treatments are used in periodontal plastic surgery to prevent and treat traumatic, structural or developmental abnormalities of the bone, alveolar mucosa, or gingiva.^[2]^ The scope for non-pocket surgical procedures has grown in significance as periodontal surgical techniques have advanced, with the objectives being amended to not only stop the disease’s progression or cure it but also to regenerate and repair the tissues that have been destroyed. In order to increase keratinized tissue, clinical attachment, and root coverage, lateral relocated flaps, free gingival grafts, coronally advanced flaps, and subepithelial connective tissue grafts have been documented over the years.^[4]^

Among the various surgical techniques employed in the management of the gingival exposure in the maxilla, the coronally advanced flap alone or combined with other graft materials have been considered as one of the most widely acceptable and predictable root coverage procedure. The chief advantage of this technique is achieving good color blend of surgically treated areas with the adjacent tissue along with the recovery of marginal soft tissue morphology.^[5]^

The denuded root surface was covered and the complete attachment apparatus was rebuilt using the guided tissue regeneration concept.^[6]^ Over time, the membranes utilized in directed tissue regeneration have changed. In order to promote regeneration, the idea of excluding the gingival epithelium with just barrier membranes has evolved to include the incorporation of the right signaling molecules, such as growth factors, and the presence of the right cell population, such as stem cells, fibroblasts, and cementoblasts, directly into the wound. One of the key cell types involved in mediating the three stages of wound healing—inflammation, proliferation, and remodeling. EGF promotes epidermal cell renewal and is crucial to the healing process for wounds, thus the application of rhEGF impregnated in collagen membrane might yield more favorable root coverage results.^[7]^ The clinical application of EGF, while fulfilling the current mechanical concept of GTR, amends it with the modern concept of biological GTR.

The goal of novel root coverage therapies is now to achieve complete root coverage with the blending of mucosa and keratinized gingiva with the adjacent tissues without any residual periodontal pocket, thereby enhancing the esthetics and reducing root sensitivity.

## MATERIALS AND METHODS

The study was designed as a randomized controlled clinical trial to evaluate the regeneration efficacy of rhEGF impregnated in absorbable collagen membrane for the management of Millers class I & class II gingival recession defects. Patients with gingival recession indicated for surgical management were recruited from the out-patient clinic, Department of Periodontics and were followed up over a period of six months after the procedure.

### Sample Size Calculation

The study would require a sample size of 15 per group calculated (total 30), for an effect size of 1.1, Probability of α error of 0.05 and a desired statistical power of 0.8, for detecting the true difference between a plain collagen membrane and rhEGF impregnated collagen membrane, in order to achieve a power of 80% and a level of significance of 5% (two sided).

### Preparation Of Recombinant Human Epidermal Growth Factor Impregnated Collagen Membrane (rhEGF)

The following steps were used to create collagen membranes containing 10 ng/ml of human recombinant epidermal growth factor (rhEGF). Type I collagen from the bovine Achilles tendon was homogenized in a 10 mm Na-butyrate solution to create standard collagen suspension (Pro Lab Marketing Pvt. Ltd, New Delhi, India). The suspension was mixed with rhEGF that had been reconstituted in 0.1M phosphate buffer. In order to cross-link collagen, 0.16% glutaraldehyde aqueous solution (Sigma Aldrich Chemicals Pvt Ltd, Bangalore, India) was added, and the resulting solution was put in separate 1.5×1.5 cm^2^ and 32 cm^2^ vats that were kept at 4°C for 12h to cross-link gelatin. In a laboratory freeze drier that is readily available, the impregnated and crosslinked membranes were frozen (Lyophilization Systems Pvt Ltd, Hyderabad, India).

### Interventions

#### Pre-Surgical Phase

After the completion of the non-surgical phase consisting of oral hygiene instructions, scaling and root surface debridement and occlusal adjustment as indicated, patients were reviewed after 4 weeks to assess the oral hygiene compliance and tissue response. After achieving satisfactory oral hygiene, patients were scheduled for the surgical procedure.

### Surgical Phase

Patients were randomly allotted, one group(test) was treated with EGF impregnated collagen membrane and the second group(control) was treated with plain collagen membrane. At the start of the surgical procedure, the patients were asked to rinse with 0.2% chlorhexidine for 1 minute.

Crevicular incisions were made from the mesial and distal aspects of the defect site after local anesthesia (Lidocaine 2%, 1:80,000 Adrenaline) was administered. Horizontal incisions were made 2 mm away from the interdental papilla connecting through intrasulcular incisions, and these were followed by reflection of a full thickness mucoperiosteal flap. The blunt dissection caused the attachments of the muscles to be severed. The interdental papillae proximal to the horizontal incisions were de-epithelialized and the root surface was debrided. Patients in the test group had an absorbable collagen membrane with rhEGF inserted on top of the recession defect, which was then stabilized with 4-0 resorbable suture material (TRUGLYDETM, Healthium Medtech Pvt. Ltd, Bangalore, India.). 4-0 non-resorbable sutures were used for tension-free suturing (TRULENETM, Healthium Medtech Pvt Ltd, Bangalore, India). COE-PAKTM, GC India Dental Pvt Ltd., a periodontal pack was placed.

In patients selected for control group, after root surface debridement, plain collagen membrane was placed on the recession defect followed by resorbable sutures. To ensure hemostasis and close adaptation of flap to the underlying surface in order to avoid dead space, gentle pressure was applied and covered using periodontal dressing.

### post-operative measures

Routine post operative instructions, antibiotics (Amoxicillin 500mg t.i.d, Metrogyl 400mg b.i.d) and analgesics (Ibuprofen 400mg + paracetamol 325mg t.i.d) for 5 days were prescribed. Patients were instructed to gently brush the area with a soft bristled tooth brush and also rinse with 0.12% chlorhexidine mouthwash twice daily. Patients were recalled after 1 week for suture removal. For recall visits, patients were reinforced with oral hygiene instructions and recalled at the end of 3-months and 6-months.

### Statistical Analysis

Version 21 of the statistical software for social sciences was used to examine the data (IBM SPSS version 21). The equality of means across groups was compared using the Independent t test, and the equality of means within groups was compared using the Paired t test. The mean scores were examined using repeated measurements at various time intervals. The p value was set at 0.05 and the confidence intervals were set at 95%.

## RESULTS

In order to treat gingival recession problems, recombinant human epidermal growth factor (rhEGF) was integrated in an absorbable collagen membrane in this prospective clinical trial. 30 isolated or numerous neighboring recession abnormalities were detected in 15 systemically healthy individuals. Following phase 1 periodontal therapy, all patients showed acceptable compliance with oral hygiene. At all the treated sites, satisfactory post-operative wound healing was seen without any untoward consequences. Additionally, patients were monitored for six months following surgery, and there were no drop-outs throughout the research period.

### Width of keratinized gingiva

The mean width of keratinized gingiva in test group were 3.47±1.36, 7.20±1.42, 7.20±1.42 and in the control group were 4.2±0.41, 5.0±0.65, 5.0±0.65 at baseline and at the end of 3- and 6-months respectively. (*Table 1 and 2*).

**Table 1:**
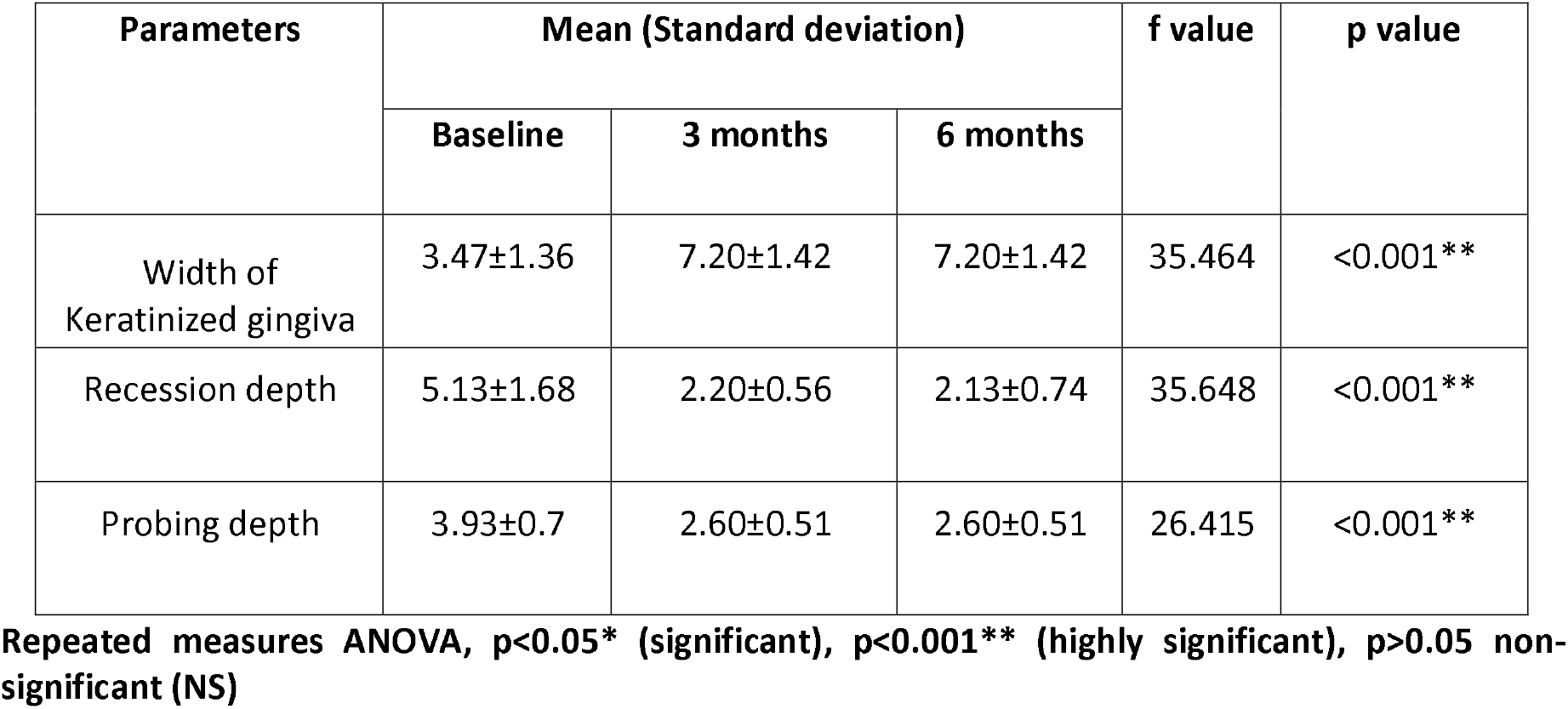
Intra group comparison of clinical parameters at base line, 3-months and 6-months in test group.

**Table 2:** Intra group comparison of clinical parameters at base line, 3 months and 6 months in control group.

| Parameters | Mean (Standard deviation) |  |  | f value | p value |
| --- | --- | --- | --- | --- | --- |
|  | Baseline | 3-months | 6-months |  |  |
| Width of Keratinized gingiva | 4.2±0.41 | 5.0±0.65 | 5.0±0.65 | 9.476 | <0.001** |
| Recession depth | 3.8±0.41 | 2.53±0.59 | 2.80±0.86 | 12.68 | <0.001** |
| Probing depth | 6.33±0.49 | 5.0±0.53 | 5.0±0.53 | 33.088 | <0.001** |

Repeated measures ANOVA, $p < 0.05^*$ (significant), $p < 0.001^{**}$ (highly significant), $p > 0.05$ non-significant (NS)

**Table 3:** Intra group comparison of clinical parameters at various intervals in test group.

| Parameters | Intervals |  | Mean difference | 95%CI |  | t value | p value |
| --- | --- | --- | --- | --- | --- | --- | --- |
|  |  |  |  | Lower limit | Upper limit |  |  |
| Width of Keratinized gingiva | Baseline | 3 months | -3.733 | -4.773 | -2.693 | -7.353 | <0.001** |
|  |  | 6 months | -3.733 | -4.773 | -2.693 | -7.353 | <0.001** |
|  | 3 months | 6 months | 0.91 | -1.06 | -0.78 | -14.22 | <0.001** |
| Recession depth | Baseline | 3 months | 2.933 | 2.16 | 3.70 | 8.191 | <0.001** |
|  |  | 6 months | 3.00 | 2.216 | 3.78 | 8.216 | <0.001** |
|  | 3 months | 6 months | 0.667 | -0.32 | 0.456 | 0.367 | <0.719* |
| Probing depth | Baseline | 3 months | 1.333 | 0.875 | 1.792 | 5.953 | <0.001** |
|  |  | 6 months | 1.333 | 0.875 | 1.792 | 5.953 | <0.001* |
|  | 3 months | 6 months | 0 | -0.379 | 0.379 | 0 | 1.00(NS) |

**Paired-t test, p<0.05\* (significant), p<0.001\*\* (highly significant), p>0.05 non-significant (NS)**

**Table 4:** Intra group comparison of clinical parameters at various intervals in control group.

| Parameters | Intervals |  | Mean difference | 95%CI |  | t value | p value |
| --- | --- | --- | --- | --- | --- | --- | --- |
|  |  |  |  | Lower limit | Upper limit |  |  |
| Width of Keratinized gingiva | Baseline | 3 months | -0.8 | -1.206 | -0 | -4.032 | <0.001** |
|  |  | 6 months | -0.8 | -1.206 | -0.394 | -4.032 | <0.001** |
|  | 3 months | 6 months | 0 | -0.486 | 0.486 | 0 | 1.00(NS) |
| Recession depth | Baseline | 3 months | 1.06 | 1.26 | 2.06 | 8.919 | <0.001** |
|  |  | 6 months | 1.00 | 0.58 | 1.41 | 5.123 | <0.001** |
|  | 3 months | 6 months | -0.67 | -1.24 | -0.08 | -2.467 | 0.027* |
| Probing depth | Baseline | 3 months | 1.33 | 0.948 | 1.712 | 7.136 | <0.001** |
|  |  | 6 months | 1.33 | 0.948 | 1.712 | 7.136 | <0.001** |
|  | 3 months | 6 months | 0 | -0.396 | 0.396 | 0 | 1.00(NS) |

**Paired-t test, p<0.05\* (significant), p<0.001\*\* (highly significant), p>0.05 non-significant (NS)**

### Recession depth

The mean of recession depth values in test group were 5.13±1.68, 2.20±0.56, 2.13±0.74 and in the control group were 3.8±0.41, 2.53±0.59, 2.80±0.86 at baseline and at the end of 3- and 6-months respectively. (*Table 1 and 2)*.

### Probing depth

At baseline and at the end of three and six months, the test group’s mean probing depth was 3.930±7, 2.600±51, 2.600±51, while in the control group it was 6.330±49, 5.00±53, 5.00±53, respectively (Table1 and 2). When comparing the baseline to the 3- and 6-month periods within the group, there are no changes in recession depth (Table 1 and 2). The mean descriptive values of probing pocket depth, recession depth, and gingival keratinization width at various time points were displayed in (Table 1). A statistically significant difference from the baseline to the 3- and 6-months was seen (p 0.05).

There was statistically significant gain in the mean values of width of keratinized gingiva from baseline to 3-months and 6-months respectively (3.47±1.36 to 7.20±1.42, p<0.001**). The gain in the recession depth from baseline to 3-months and 6-months was also statistically significant (5.13±1.68 to 7.20±1.42, p<0.001**). The mean descriptive values of pocket probing depth were 3.93±0.7, 2.60±0.51, 2.60±0.51, p<0.001**). All the parameters did not show any significant changes from 3-months to 6-months.

Statistically highly significant difference was observed from baseline to 3-months and significant difference from 3-months to 6-months between groups for width of keratinized gingiva at different intervals (*Table 5*). No significant difference was observed from baseline to 3-months and significant difference was observed from baseline to 6-months for recession depth at different intervals in between the group. Highly significant was observed from baseline to 3-months for probing depth at different intervals in between the group (*Table 5*).

**Table 5:** Inter group comparison of clinical parameters at various intervals in test and control group.

| Variable | Interval | Test | Control | t-value | P value |
| --- | --- | --- | --- | --- | --- |
| Width of Keratinized gingiva | Base line | 3.47±1.36 | 4.2±0.41 | -2.004 | 0.055* |
|  | 3 months | 7.20±1.42 | 5.0±0.65 | 5.435 | 0.000** |
|  | 6 months | 7.20±1.42 | 5.0±0.53 | 5.435 | 0.000** |
| Recession depth | Base line | 5.13±1.68 | 3.8±0.41 | 2.977 | 0.006 |
|  | 3 months | 2.20±0.56 | 2.73±0.59 | -2.53 | 0.017** |
|  | 6 months | 2.13±0.74 | 2.8±0.86 | -2.269 | 0.033* |
| Probing depth | Base line | 3.93±0.7 | 6.33±0.49 | -10.854 | 0.001** |
|  | 3 months | 2.60±0.51 | 5.0±0.53 | -12.616 | 0.001** |
|  | 6 months | 2.60±0.51 | 2.27±0.59 | -12.616 | 0.001** |

**Independent t test, p<0.05\* (significant), p<0.001\*\* (highly significant), p>0.05 non-significant (NS)**

## DISCUSSION

In the early years following the development of mucogingival surgery, procedures were primarily intended to improve the relationship between the gingiva and the alveolar mucous membrane, with issues like inadequately attached gingiva, a shallow vestibule, and a high frenal attachment being the main focus. The goal of recession management was to stop the recession from getting worse while improving biofilm control and retaining keratinized tissue. [8] In class I and class II gingival recession sites, the current study showed the adjunctive effect of rhEGF impregnated collagen membrane and compared its efficacy to plain collagen membrane.

EGF acts as a versatile regulator of wound healing and plays a role in angiogenesis providing beneficial effects in regenerative therapy.^[9]^ Collagen shows properties such as hemostatic activity, attracts and activates neutrophils, fibroblasts and also interacts with various cells during tissue re-modelling.^[10]^ Its low immunogenicity made collagen an attractive biomaterial. When these two materials are used together it showed synergistic effects revealing the beneficial effect of this combination.

Every stage of the cascade of wound healing requires collagen. Collagen has been utilized successfully in the treatment of burns, non-healing ulcers, traumatic and surgical wounds, and different aesthetic operations as a local medication delivery system.^[11]^ The short half-life of rhEGF, which results from its oxidation by the hydrolytic enzymes present in the wound bed, makes it difficult to use in clinical settings. Collagen matrices employed in soft tissue augmentations have demonstrated sufficient volume stability, giving cells enough time to infiltrate the collagen matrix and regenerate new soft tissue.^[12]^ Because of this, type I collagen was utilized in the current study as a scaffold to delay the release of EGF and the collagen membrane was crosslinked with 0.16% of glutaraldehyde aqueous solution in order to delay the degradation of the membrane.

In the present study the width of keratinized gingiva showed a significant increase from baseline to 3-months and showed constant results from 3-months to 6-months (p≤0.001). These findings are in accordance with the observations^[13]^ where PRF was used to increase wKG due to growth factors release from the fibrin mesh, thereby influencing the proliferation and migration of gingival and periodontal fibroblasts, leading to the formation of connective tissue which increases keratinization of the overlying epithelium.

Fibroblasts are one of the various cell types that actively contribute to the process of tissue repair by multiplying, migrating and filling the wound in addition to helping to produce growth factors and extracellular matrix components. EGF plays a role in fibroblasts, particularly in the process of re-epithelialization by enhancing collagen formation and promoting cell renewal.^[14]^ However, EGF’s proteolytic nature makes it easily destroyed in the sites of wounds. There are other reports that fibrin or fibrinogen can increase these processes. With the use of these information, it can be concluded that rhEGF are biologically safe and can be used to encourage the regeneration of soft tissue and improve wound healing.

In the present study, in control group on comparing the width of keratinized gingiva from base line to 6-months a significant amount of increase in keratinized gingiva was noted (p=0.001), but there was not much difference seen from 3-months to 6-months (p=0.001). Attachment outcome results with GTR reported in the literature showed consistently significant formation of bone, cementum and connective tissue attachment.

In GTR operations, the proliferation of granulation tissue, which was able to promote keratinization, results in a moderate increase in the width of keratinized tissue.^[15]^ As a result, a notable decrease in PPD was observed from baseline to three months, and it remained consistent from three months to six months, with a mean increase of 1.33mm.

Overall, on inter group comparison, the wKG when compared from baseline to 3-months, there was statistically significant increase in both the groups (p<0.001). There was no noticeable variation from 3-months to 6-months in both the groups which showed comparable results. This increase in wKG was may be due to fibrin linkage, which may facilitate initial clot formation and stabilization, leading to enhanced regeneration.^[16]^

A mean increase in gingival thickness of 0.070±03mm and 0.300±10mm respectively, was observed by Gupta and Thamaraiselvan et al., Because rhEGF contains bioactive proteins that support neoangiogenesis and tissue ingrowth, the considerable increase in gingival thickness can be attributed to these factors. According to research by Aroca et al., Cardarapoli et al., Stefanini et al., Santosh gupta et al., and Atilla et al., the usage of collagen scaffolds improved gingival thickness. The gingival thickness has increased in the current study in both the test group and control group, similarly.^[17-22]^

In the present study, all the patients had very minimal PPD of 1-2mm only, at baseline. No changes in PPD were noted at all time intervals in both the groups (p=0.001). In test sites no similar comparisons could be made due to lack of evidences using rhEGF in recession defects but it could be expected that addition of growth factors could result in attaining better clinical outcomes.

In control site the mean PD reduction from baseline to 6-months was 6.33±0.49 mm to 2.27±0.59 mm and a mean reduction of 1.33mm was achieved in the present study. The mean PD at the baseline was 3.93±0.7 mm which significantly reduced to 2.60±0.51mm in test sites with mean reduction of 1.3 mm. The control site showed better outcomes in terms of PPD but the difference was not statistically significant. At 3-months and 6-months PPD showed statistically significant reduction (p≤0.05). Between 3-months and 6-months there was no significant change in PD demonstrating the stability of results achieved. The decreased reduction in PD in test and control group could be attributed to slightly increased mean at baseline in test sites when compared to that of control sites though the difference was not statistically significant.

In the present study RD showed statistically significant reduction at baseline and 3-months (p≤0.05) in both test and control groups, and showed increase from 3-months to 6-months in control group, further from 3-months to 6-months in test group there was statistically significant change in the RD reduction. It might be speculated that creeping attachment over the natural teeth might be more predominant due to the positive and favourable cellularity provided by the periosteum and the capacity of the periodontal ligament to proliferate over a denuded root surface.

To best of our knowledge this is the first human clinical trial on the usage of rhEGF impregnated collagen membrane in Miller’s Class 1 and 2 for gingival recession defects. At the end of 6-months follow up, in the current study we observed significant improvement in terms of increase in width of keratinized gingiva, probing depth and recession depth with limited complications, suggesting the therapeutic benefits of rhEGF application in managing Miller’s Class 1 and 2 for gingival recession defects. Considering the promising results in the present study, this prospective clincal study was therefore performed in order to ascertain the efficacy of rhEGF impregnated collagen membrane in Miller’s Class 1 and 2 for gingival recession defects. However, studies on the efficacy of GTR have shown predictability with use of a collagen membrane. With the cumulative observations made in the present study it can be proposed that rhEGF impregnated collagen membrane showed proper wound healing, keratinocytes proliferation and fibroblast migration.

The potential limitations of this study are histologic examination of the tissue was not evaluated; gingival biotype was not assessed. Further studies with a larger sample size and extended periods of follow up are required to explore the synergistic effects of rhEGF in various domains of periodontal plastic surgery.

## CONCLUSION

A comparative clinical study was done to assess the efficacy of rhEGF impregnated collagen membrane over plain collagen membrane in Millers class I and class II gingival recession defects. Following conclusions were made with reference to the observation in this study.

1. rhEGF impregnated collagen membrane provided added benefits over collagen membrane alone, when various parameters were compared.
2. In gingival recession defects where root coverage was performed using rhEGF impregnated collagen membrane, there was a sufficient gain in width of keratinized gingiva, reduction in recession depth and improvement in probing depths.

## LEGENDS OF FIGURES

**Figure-1.**
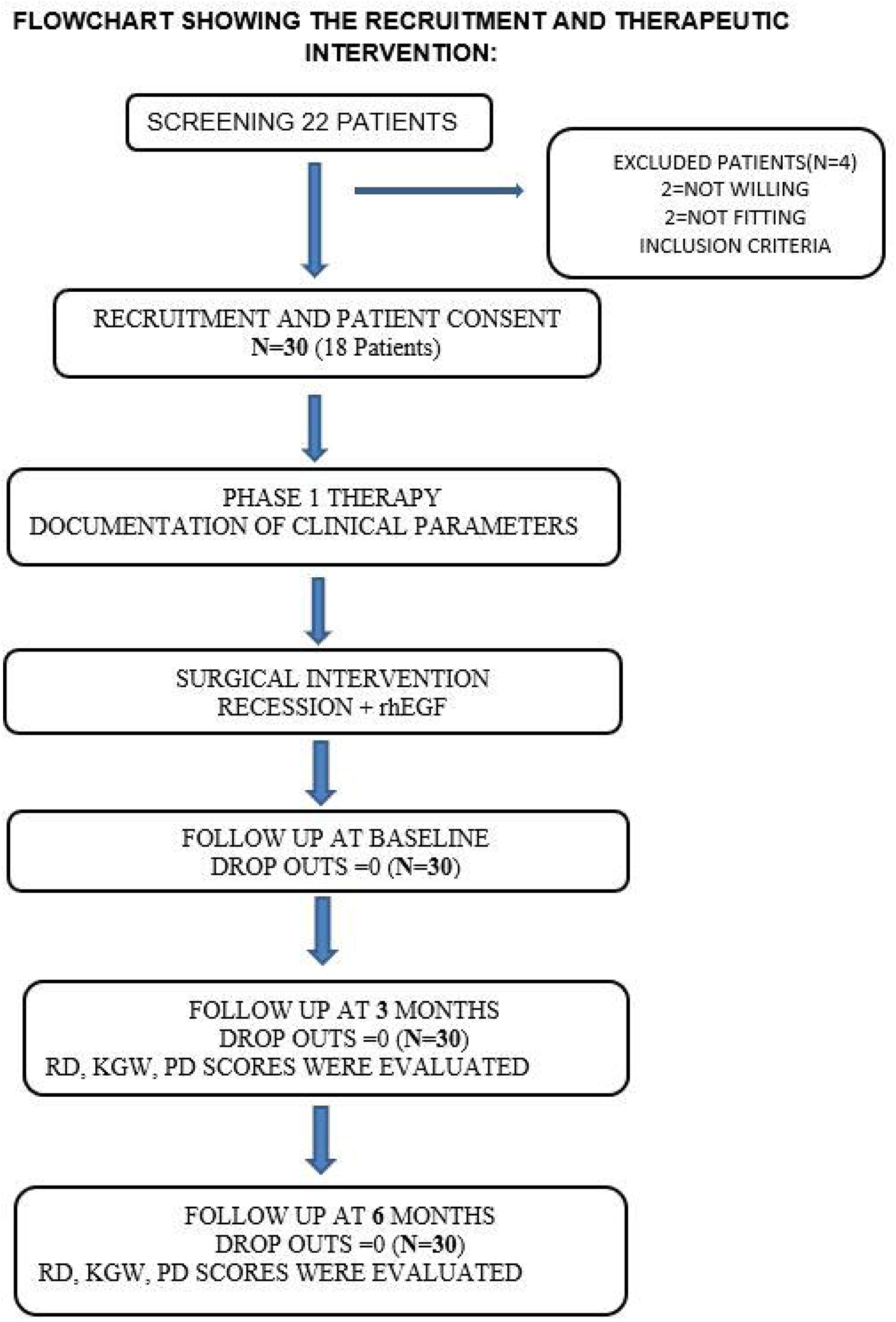
Flowchart showing the recruitment and therapeutic intervention

**Figure-2.**
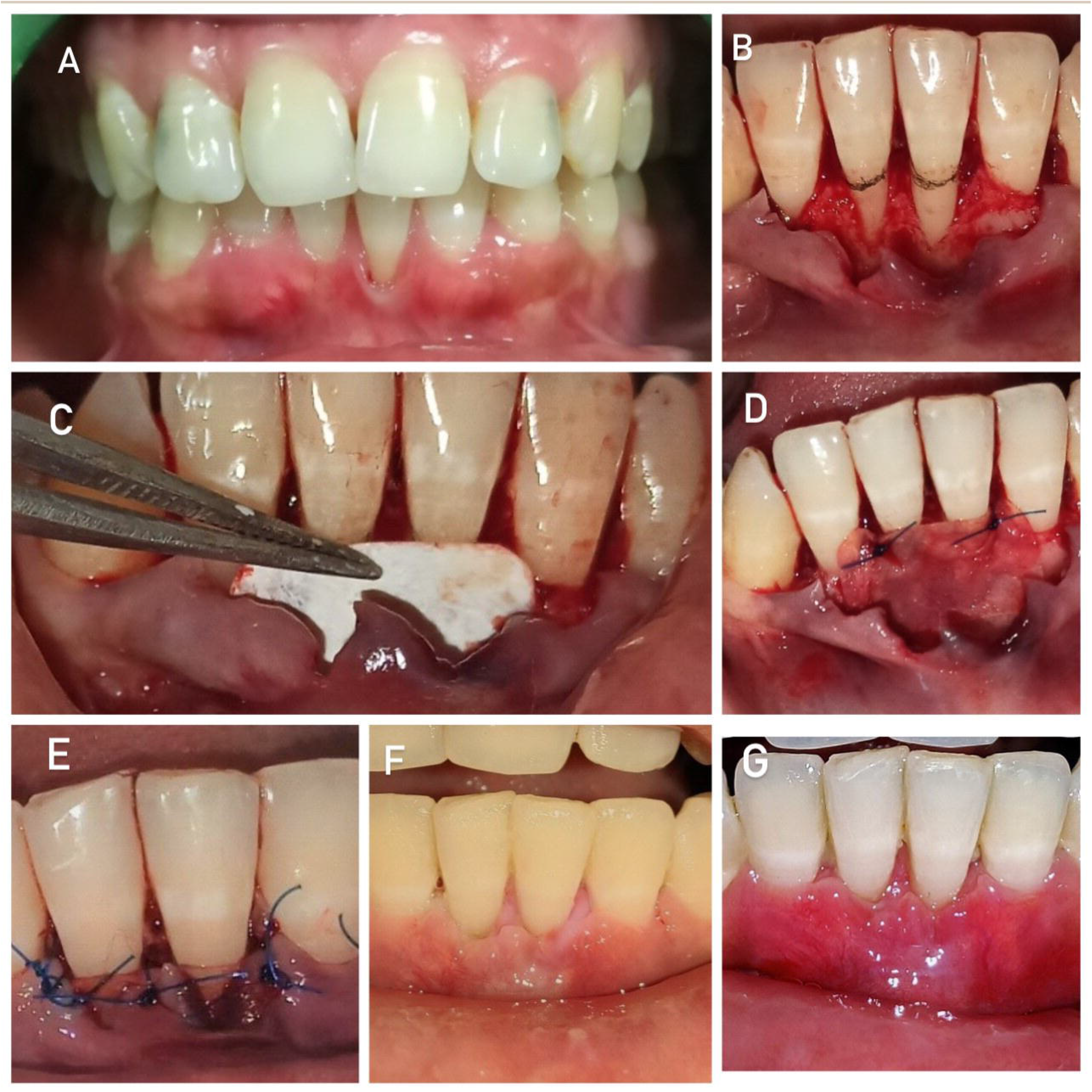
**2a)** Millers class 1 gingival recession irt 31,41; **2b)** Full thickness mucoperiosteal flap was reflected followed by thorough debridement of the site. de-epithelization of the papillae was done; **2c)** rhEGF impregnated collagen membrane was placed upon the recession site; **2d)** rhEGF impregnated collagen membrane was stabilized with 4-0 resorbable sutures; **2e)** flap was approximated and 4-0 non-resorbable sutures were placed; 2f) 1-week post-operative photographs; 2g) 1-month post-operative photographs

**Figure-3.**
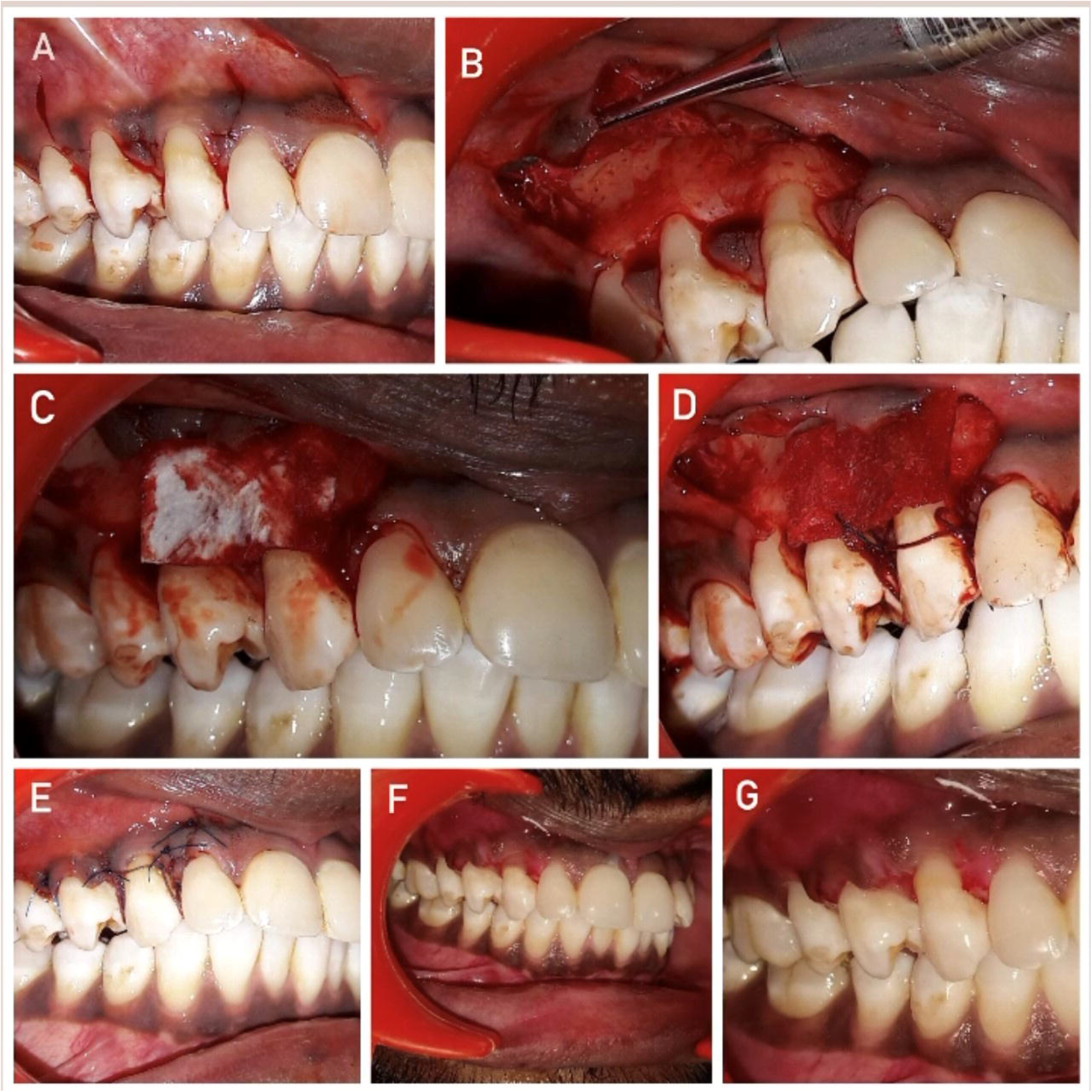
**3a)** Millers class ii gingival recession irt 13,14; **3b)** Horizontal incisions were given at mesial and distal aspects of involved tooth at the base of the papillae followed by crevicular & two vertical releasing incisions; **3c)** Full thickness mucoperiosteal flap was reflected followed by thorough debridement of the site. de-epithelization of the papillae was done; **3d)** Collagen membrane was placed upon the recession site; **3e)** Membrane was stabilized with 4-0 resorbable suture material; **3f)** Flap was approximated and 4-0 non-resorbable sutures were placed; 3g) 1week post-operative photographs; 3h) 1-month post-operative photographs

**Figure-4.**
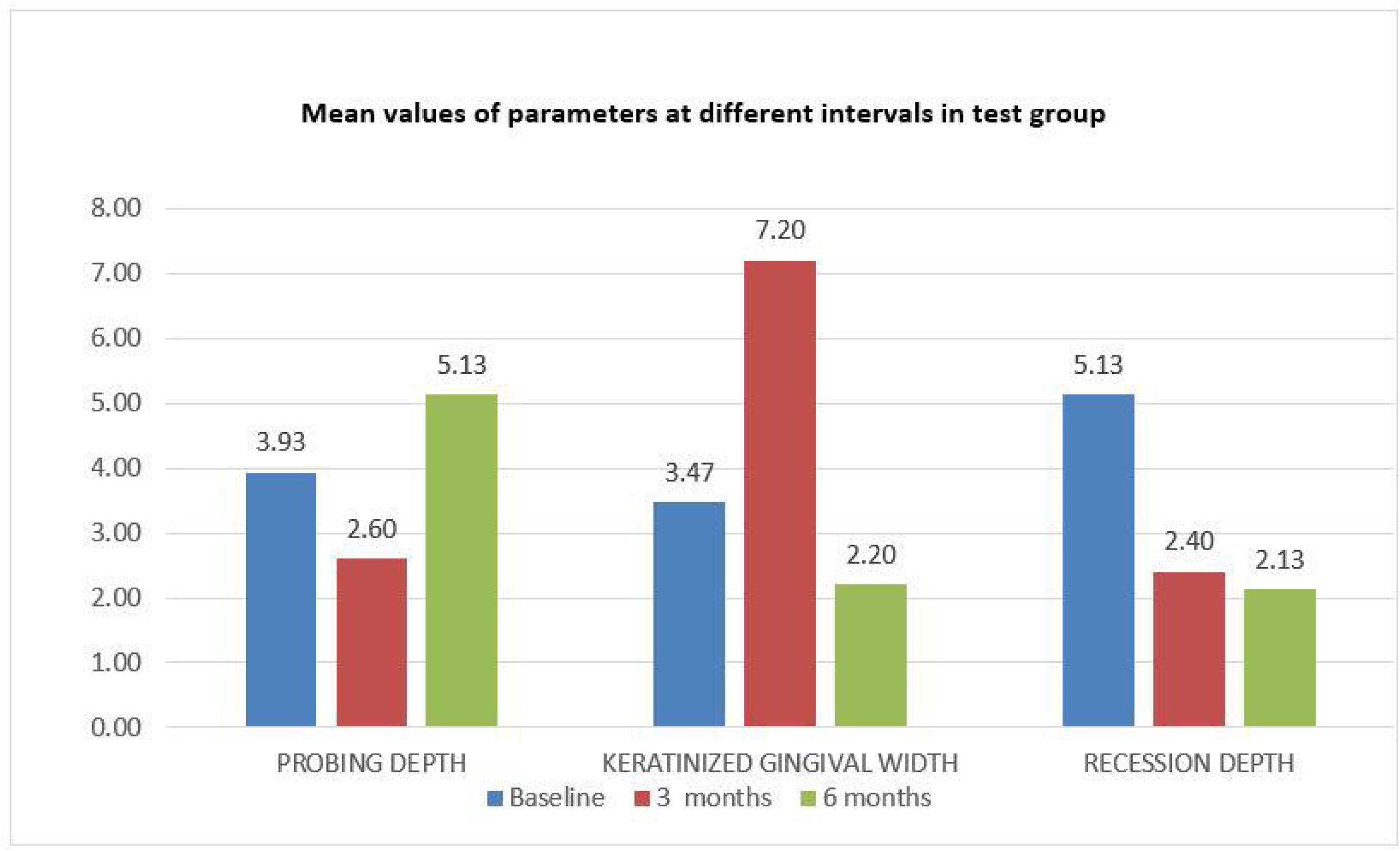
Mean values of parameters at different intervals in test group

**Figure-5.**
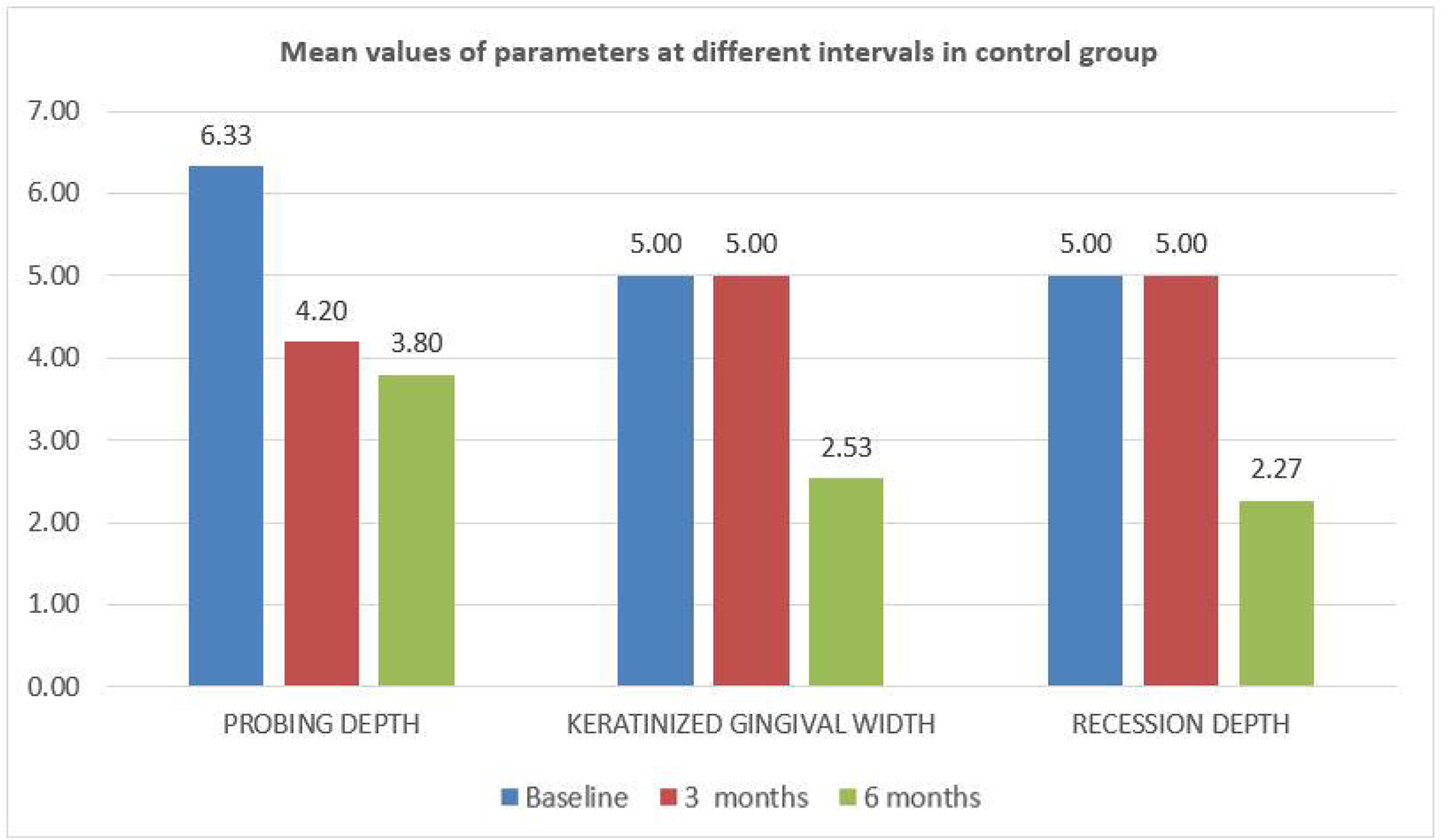
Mean values of parameters at different intervals in control group

**Figure-6.**
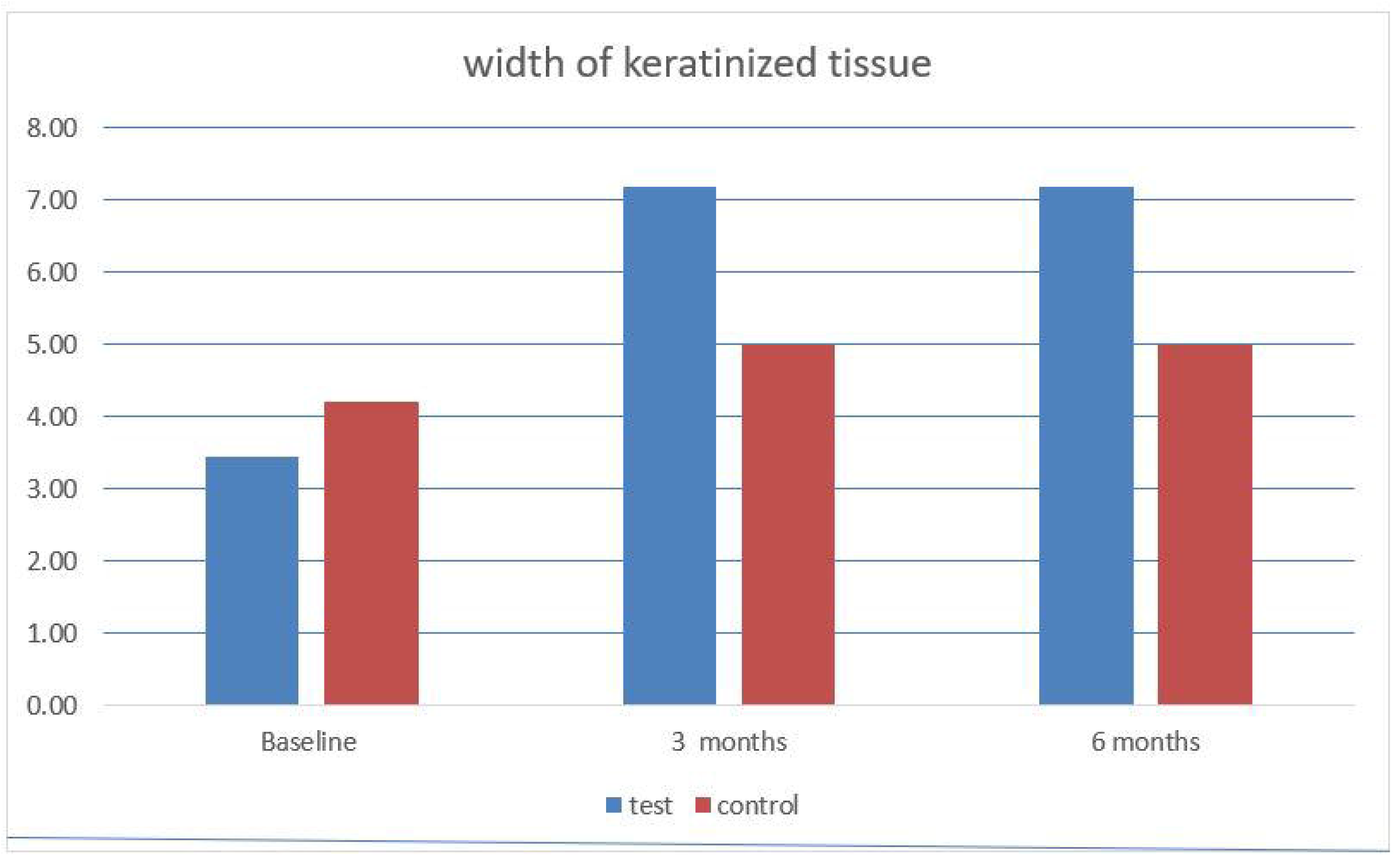
Mean values of width of keratinized gingiva at different interval in test and control group

**Figure-7.**
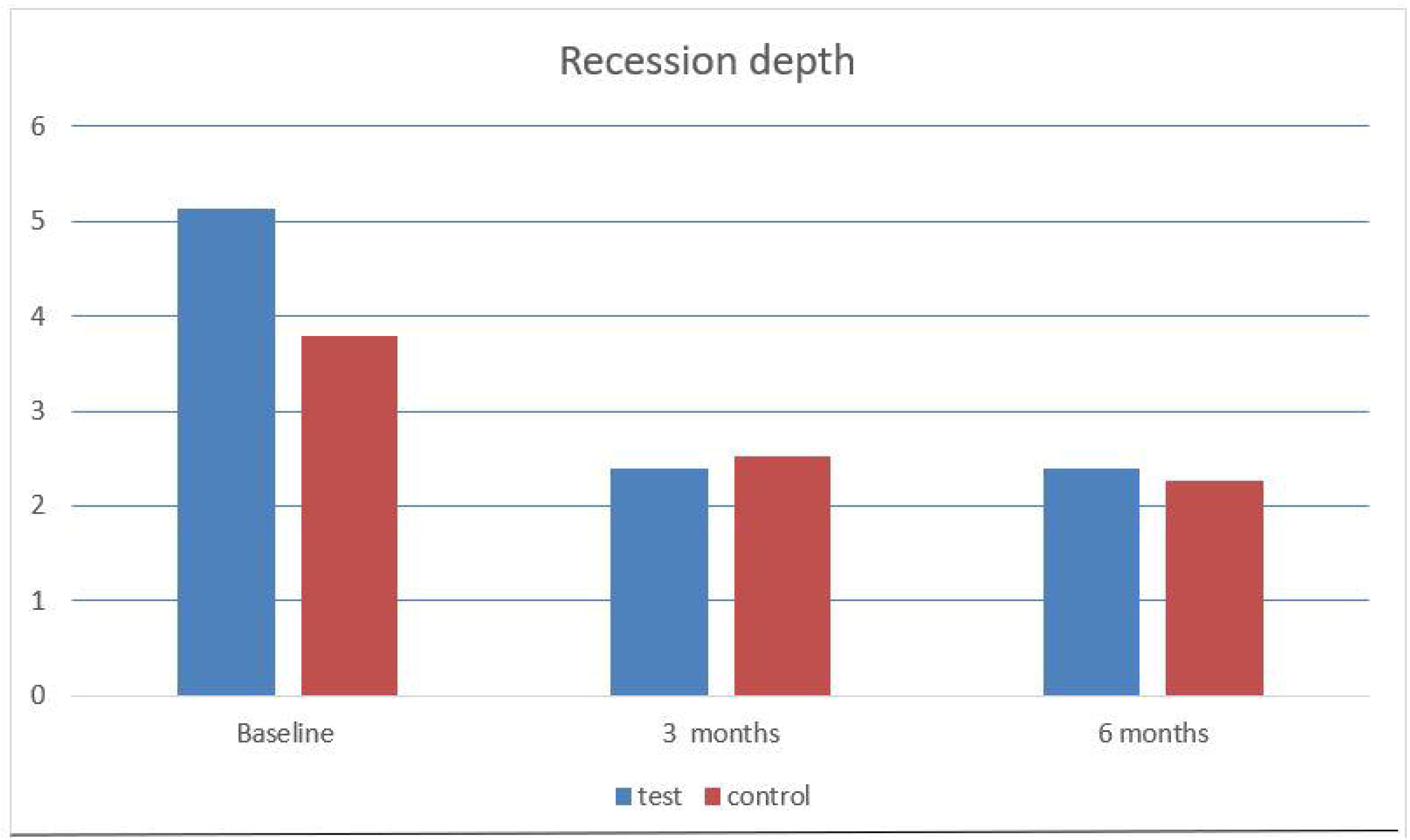
Mean values of recession depth at different intervals in test and control group

**Figure-8.**
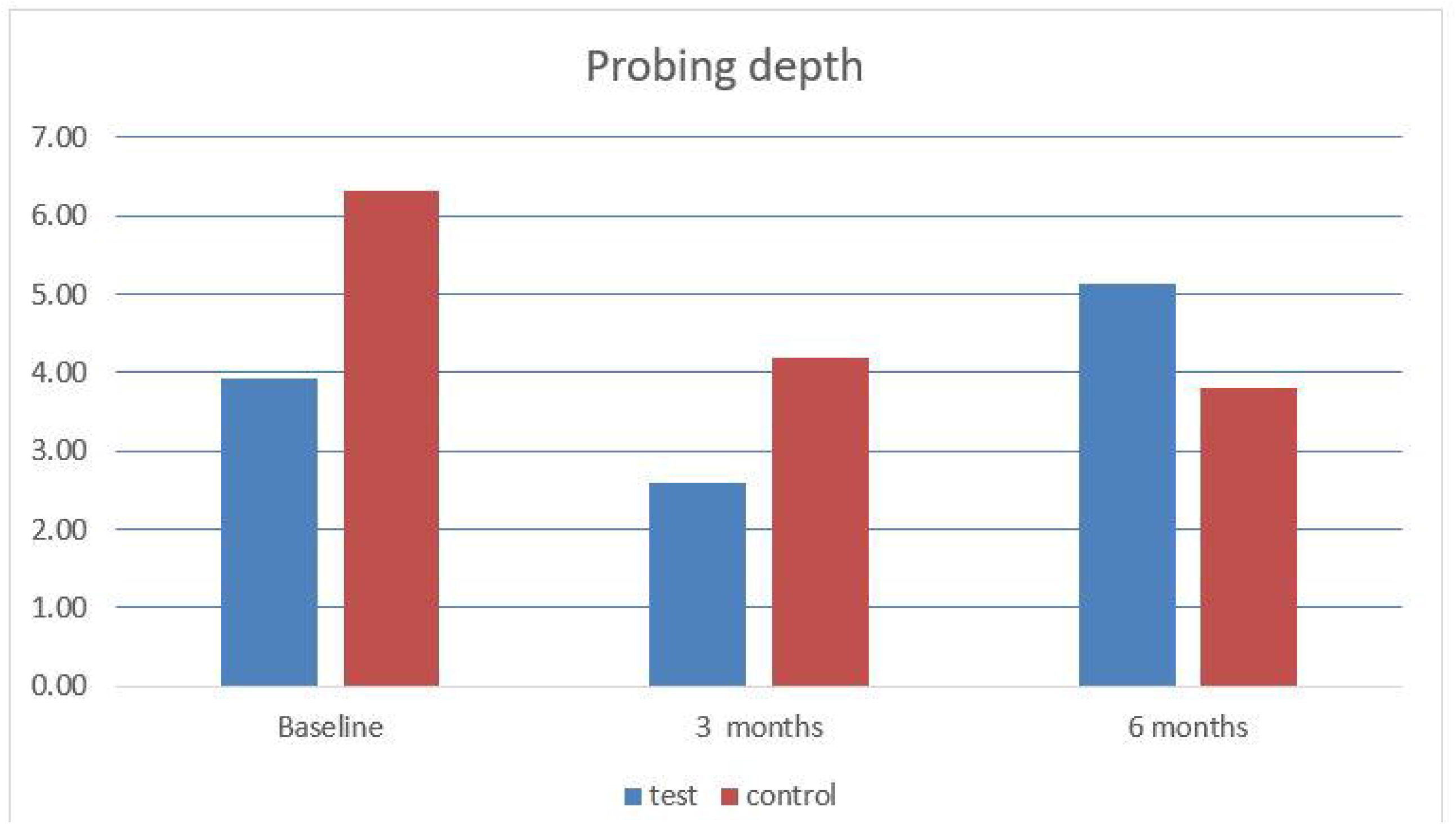
Mean values of probing depth of gingiva at different intervals in test and control group

## Data Availability

All data produced in the present work are contained in the manuscript

